# 2024-2025 BNT162b2 COVID-19 vaccine effectiveness in non-immunocompromised adults: mid-season estimates from vaccine registries in two states linked to administrative claims

**DOI:** 10.1101/2025.05.28.25327615

**Authors:** Kathleen M. Andersen, Tara Ahi, Jazmine S. Mateus, Tiange Yu, Anan Zhou, Santiago M.C. Lopez, Laura Puzniak

## Abstract

**Background:** Data are limited on 2024-2025 BNT162b2 COVID-19 vaccine effectiveness (VE).

**Methods:** Retrospective cohort study among non-immunocompromised adults from August 22, 2024 (“index”) to December 31, 2024, among residents of California or Louisiana continuously enrolled in health insurance plans reporting to HealthVerity for ≥1 year prior to index. Receipt of 2024-2025 BNT162b2 COVID-19 vaccine was defined using state vaccine registries with health insurance claims, using a time-varying exposure definition. VE against COVID-19-associated hospital admissions was estimated as (1-hazard ratio), using adjusted Cox proportional hazards models with 95% confidence intervals (CI).

**Results:** Overall, 6,900,361 individuals met selection criteria for the study. By the end of follow-up (median 4.4 months), 325,362 (4.7%) had received a BNT162b2 2024-2025 COVID-19 vaccine dose. VE against COVID-19-associated hospital admission was 41% (95% CI 2-64).

**Discussion:** The 2024-2025 formulation of BNT162b2 COVID-19 vaccine provided significant protection, particularly for older adults, in mid-season estimates.

This study is registered on clinicaltrials.gov as NCT06923137.

## INTRODUCTION

On June 27, 2024, the Centers for Disease Control and Prevention (CDC) Advisory Committee on Immunization Practices (ACIP) recommended 2024–2025 COVID-19 vaccination with an FDA–authorized or approved vaccine for all persons aged ≥6 months.^1^ This recommendation aligns with past regulatory decisions to update formulation based on the antigenic divergence of the SARS-CoV-2 virus in an effort to help prevent severe outcomes of infection including COVID-19-associated hospitalization and death.^2^ On August 22, 2024, the United States (US) Food and Drug Administration (FDA) approved an updated monovalent messenger RNA (mRNA) COVID-19 vaccine formulation targeting the Omicron KP.2 strain for individuals 12 years and older and authorized for individuals ≥6 months to 12 years.^3^

While prior studies have described the vaccine effectiveness (VE) of mRNA COVID-19 vaccines, there are limited temporal data on 2024-2025 COVID-19 vaccines and estimates by age groups.^4,5^ These data are critical to inform public health policy regarding future COVID-19 vaccine recommendations and guide clinicians and public stakeholders. This study uses a novel approach to maximize the reliability of vaccination reporting by combining state vaccine registries with mandatory reporting and linking administrative claims data. We estimated mid-season 2024-2025 BNT162b2 COVID-19 VE against COVID-19-associated hospital admission by various age groups in two heterogenous states in the US.

## METHODS

### Design, Setting, and Participants

We conducted a retrospective cohort study among non-immunocompromised adults aged 18 years and older beginning August 22, 2024 (“index date”, date of FDA approval of BNT162b2 2024-2025 season formulation targeting the SARS-COV-2 Omicron KP.2 sub-variant).^3^ Individuals were followed from index date until the outcome of interest (described below), or were censored at the first of December 31, 2024, disenrollment from medical and/or pharmacy plans, receipt of a non-BNT162b2 vaccine, or receipt of a second 2024-2025 dose of any kind after receiving a first 2024-2025 BNT162b2 dose. Details on the methodology of the study design have been previously described.^6–8^ In brief, the study was limited to individuals without pharmacologic or diagnostic evidence of immunocompromised status who were residents of California or Louisiana and continuously enrolled in health insurance plans reporting to HealthVerity for at least one year prior to index.^9^ Individuals with missing information for age and/or sex were excluded, as were people who had a documented case of COVID-19 and/or a COVID-19 vaccine in the 0-90 days prior to index. This study utilized deidentified data, was reviewed by the Sterling Institutional Review Board (ID #13433), and deemed exempt from further ethical oversight. The study protocol was posted on clinicaltrials.gov (NCT06923137) prior to initiation of all analyses.

### Exposure

Non-immunocompromised adults who received the BNT162b2 2024-2025 formulation were compared to non-immunocompromised adults who had not received any 2024-2025 season COVID-19 vaccine. Vaccination was treated as a time-varying exposure, with individuals censored from unvaccinated person-time on the date of vaccination and considered vaccinated 14 days after receiving a 2024-2025 BNT162b2 COVID-19 vaccine to reflect sufficient time to develop antibody protection.^6,8^

As previously described, any record of COVID-19 vaccine in state vaccine registries via Clinical Vaccines Administered (CVX) code, pharmacy claims using National Drug Codes (NDC), and/or medical claims using Current Procedural Terminology (CPT) codes were considered.^6–8^ Person-time in which individuals did not have a record of receiving any brand of a 2024-2025 COVID-19 vaccine in both the state registry and claims data was categorized as unvaccinated.

### Outcomes

COVID-19-associated hospitalization was defined using International Classification of Diseases, Tenth Revision, Clinical Modification (ICD-10-CM) code U07.1 “COVID-19” for inpatient admission. Hospitalizations were excluded if there were any additional ICD-10-CM codes for the same admission indicating that COVID-19 was an incidental finding concomitant to unintentional injury, physical trauma, poisoning, short-stay childbirth, or severe and persistent mental illness.^6,10^ Hospitalization was modeled separately for vaccinated and unvaccinated periods at risk and by age (≥18 years, ≥50 years and ≥ 65 years.

### Statistical Analyses

Categorical variables are reported as counts with percentages (%), and continuous variables are reported as mean with standard deviation (SD) or median with first and third quartiles (Q1, Q3). Standardized mean differences (SMD) were used to compare descriptive statistics in a sample size independent manner, with >0.1 indicating imbalance between vaccinated and unvaccinated groups. Incidence rates are shown as the number of events per 100,000 person-months, separately for vaccinated and unvaccinated time. Cox proportional hazards models were used to estimate hazard ratios (HR), sex (male, female), state of residence (California or Louisiana), insurance payor (Medicaid, Medicare, commercial), at least one CDC-defined high-risk condition for severe COVID-19 (presence, absence), wellness visit in year prior to index (presence, absence), influenza vaccination in year prior to index (presence, absence), outpatient encounter in 180 days prior to index (presence, absence), ED encounter in 180 days prior to index (presence, absence), and medically-attended COVID-19 in the 91 to 365 days prior to index (presence, absence).^6,11–13^ In the models not stratified by age, we additionally adjusted for age (18-64, ≥65 years of age). We calculated 95% confidence intervals using the sandwich estimator, given individuals who were vaccinated had non-independent repeated measures in the time-varying exposure framework, and with the Efron method to address ties in times-to-event, given the compressed follow-up time.^14^

Vaccine effectiveness (VE) was calculated as (1 – adjusted HR), multiplied by 100 to create a percentage, and can be interpreted as the incremental benefit of receiving the 2024-2025 BNT162b2 vaccine in a population with a high level of pre-existing immunity from prior infection, vaccination or a combination of the two.^5^ Analyses were conducted with one programmer using SAS, version 9.4 (SAS Institute Inc.) and a second programmer using R, version 4.5.0 (R Foundation for Statistical Computing) to assure reliability of results.

## RESULTS

### Cohort descriptives

A total of 6,900,361 persons met selection criteria for the study (6,402,993 [93%] from California and 497,368 [7%] from Louisiana; 2,801,573 [41%] were aged ≥50 years and 1,128,493 [16%] ≥65 years). The median follow-up time was 4.4 months (Q1: 4.4, Q3: 4.4) and did not differ substantially by state or age. By the end of the study, 325,362 (4.7%) received the BNT162b2 2024-2025 COVID-19 vaccine; 316,693 (4.9%) in California and 8,669 (1.7%) in Louisiana.

Compared to those who had not received a 2024-2025 season COVID-19 vaccine of any brand, the 2024-2025 BNT162b2 vaccinated group tended to be older (mean age 54 vs 45 years, SMD: 0.52) **(Table 1)**. Vaccinated individuals were more likely to have commercial insurance (59% vs 41%, SMD: 0.65), a wellness visit (46% vs 33%, SMD: 0.29), influenza vaccination (62% vs 17%, SMD = 1.05) in the prior year, ever received a pneumococcal (21% vs 7%, SMD = 0.39) or herpes zoster (30% vs 9%, SMD = 0.56) vaccine, and an outpatient encounter in the prior 180 days (70% vs 53%, SMD = 0.35). Additionally, vaccinated individuals were more likely to have received any brand of the 2023-2024 XBB.1.5-adapted COVID-19 vaccine (65% vs 9%, SMD: 1.44).

**Table 1:**
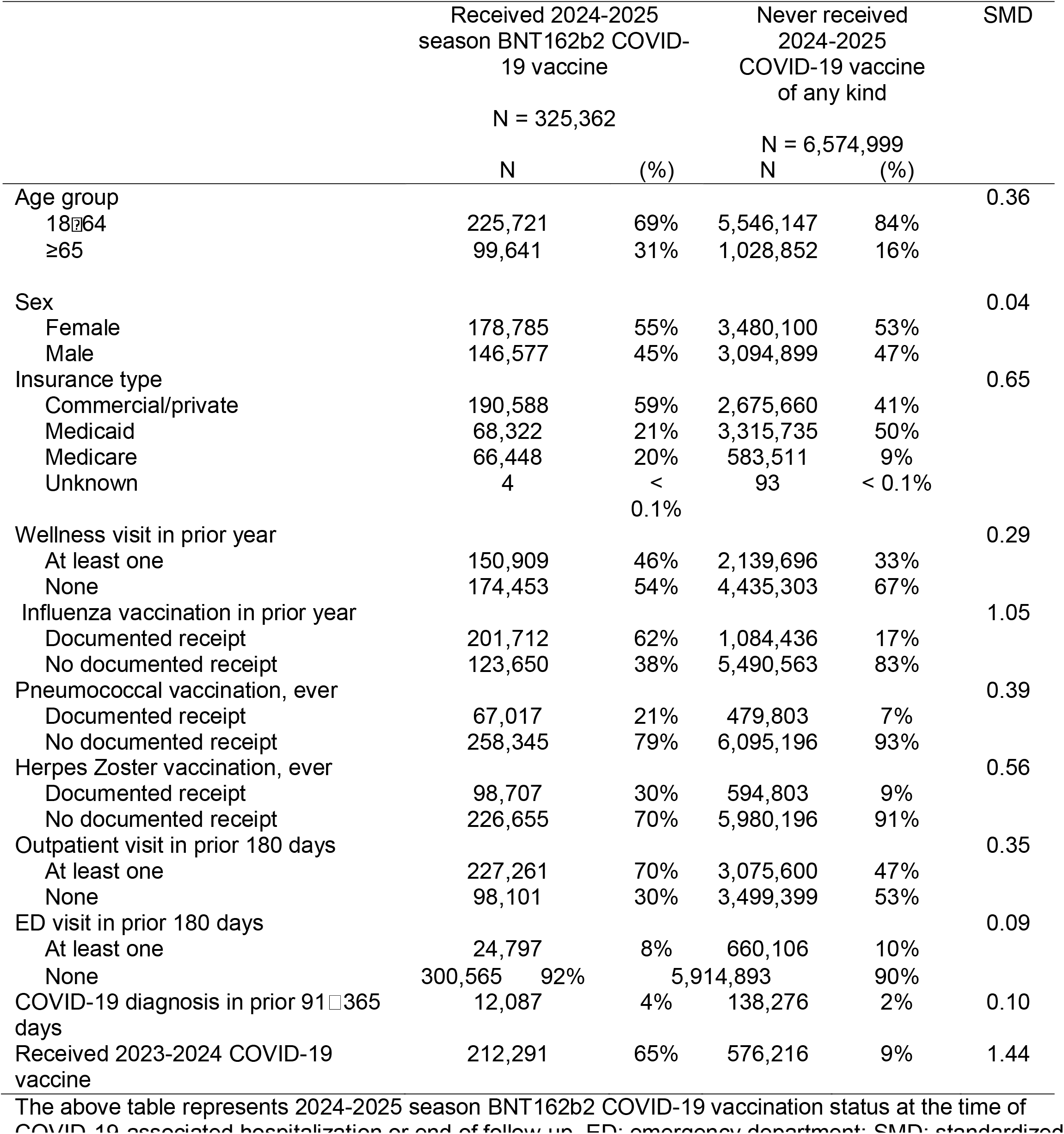
Cohort Descriptives.

|  | Received 2024-2025<br>season BNT162b2 COVID-19 vaccine |  | Never received<br>2024-2025<br>COVID-19 vaccine<br>of any kind |  | SMD |
| --- | --- | --- | --- | --- | --- |
|  | N = 325,362 |  | N = 6,574,999 |  |  |
|  | N | (%) | N | (%) |  |
| Age group |  |  |  |  | 0.36 |
| 18-64 | 225,721 | 69% | 5,546,147 | 84% |  |
| ≥65 | 99,641 | 31% | 1,028,852 | 16% |  |
| Sex |  |  |  |  | 0.04 |
| Female | 178,785 | 55% | 3,480,100 | 53% |  |
| Male | 146,577 | 45% | 3,094,899 | 47% |  |
| Insurance type |  |  |  |  | 0.65 |
| Commercial/private | 190,588 | 59% | 2,675,660 | 41% |  |
| Medicaid | 68,322 | 21% | 3,315,735 | 50% |  |
| Medicare | 66,448 | 20% | 583,511 | 9% |  |
| Unknown | 4 | < 0.1% | 93 | < 0.1% |  |
| Wellness visit in prior year |  |  |  |  | 0.29 |
| At least one | 150,909 | 46% | 2,139,696 | 33% |  |
| None | 174,453 | 54% | 4,435,303 | 67% |  |
| Influenza vaccination in prior year |  |  |  |  | 1.05 |
| Documented receipt | 201,712 | 62% | 1,084,436 | 17% |  |
| No documented receipt | 123,650 | 38% | 5,490,563 | 83% |  |
| Pneumococcal vaccination, ever |  |  |  |  | 0.39 |
| Documented receipt | 67,017 | 21% | 479,803 | 7% |  |
| No documented receipt | 258,345 | 79% | 6,095,196 | 93% |  |
| Herpes Zoster vaccination, ever |  |  |  |  | 0.56 |
| Documented receipt | 98,707 | 30% | 594,803 | 9% |  |
| No documented receipt | 226,655 | 70% | 5,980,196 | 91% |  |
| Outpatient visit in prior 180 days |  |  |  |  | 0.35 |
| At least one | 227,261 | 70% | 3,075,600 | 47% |  |
| None | 98,101 | 30% | 3,499,399 | 53% |  |
| ED visit in prior 180 days |  |  |  |  | 0.09 |
| At least one | 24,797 | 8% | 660,106 | 10% |  |
| None | 300,565 | 92% | 5,914,893 | 90% |  |
| COVID-19 diagnosis in prior 91-365 days | 12,087 | 4% | 138,276 | 2% | 0.10 |
| Received 2023-2024 COVID-19 vaccine | 212,291 | 65% | 576,216 | 9% | 1.44 |

### Prevalence of high-risk conditions

CDC-defined high-risk conditions were common in all individuals but were more prevalent in the vaccinated group (70% vs 61%, SMD: 0.2) for the twenty comorbidities evaluated, compared to the unvaccinated (**Table 2)**. The most common comorbid conditions overall were obesity, mental health conditions, heart conditions, cancer and type 2 diabetes mellitus.

**Table 2:** Prevalence of High-Risk Conditions Conferring Increased Risk of Severe COVID-19.

|  | Received 2024-2025<br>season BNT162b2<br>COVID-19 vaccine |  | Never received<br>2024-2025<br>COVID-19 vaccine<br>of any kind |  | SMD |
| --- | --- | --- | --- | --- | --- |
|  | N = 325,362 |  | N = 6,574,999 |  |  |
|  | N | % | N | % |  |
| At least one of the following conditions | 228,430 | 70% | 3,991,261 | 61% | 0.20 |
| Obesity | 88,152 | 27% | 1,765,281 | 27% | 0.01 |
| Mental health conditions | 84,424 | 26% | 1,437,939 | 22% | 0.10 |
| Heart conditions | 80,908 | 25% | 1,045,852 | 16% | 0.22 |
| Cancer | 77,506 | 24% | 834,719 | 13% | 0.29 |
| Diabetes mellitus, type 2 | 70,508 | 22% | 1,012,410 | 15% | 0.16 |
| Disabilities, including Down syndrome | 50,924 | 16% | 862,668 | 13% | 0.07 |
| Cerebrovascular disease | 43,478 | 13% | 509,428 | 8% | 0.18 |
| Smoking, current and former | 40,234 | 12% | 792,925 | 12% | 0.01 |
| Chronic kidney disease | 34,071 | 10% | 373,615 | 6% | 0.18 |
| Chronic lung diseases | 32,757 | 10% | 432,100 | 7% | 0.13 |
| Chronic liver diseases | 27,967 | 9% | 474,658 | 7% | 0.05 |
| Primary immunodeficiencies | 8,961 | 3% | 85,984 | 1% | 0.10 |
| Asthma | 7,301 | 2% | 103,494 | 2% | 0.05 |
| Use of corticosteroids or other immunosuppressives | 6,361 | 2% | 82,505 | 1% | 0.06 |
| Diabetes mellitus, type 1 | 5,020 | 2% | 75,623 | 1% | 0.03 |
| Tuberculosis | 4,742 | 1% | 89,117 | 1% | 0.01 |
| Neurologic conditions | 2,537 | 1% | 32,106 | 0.5% | 0.04 |
| HIV | 1,080 | 0.3% | 16,904 | 0.3% | 0.01 |
| Solid organ or blood stem transplantation | 466 | 0.1% | 6,318 | 0.1% | 0.01 |
| Cystic fibrosis | 121 | < 0.1% | 3,170 | < 0.1% | 0.01 |

### COVID-19-associated hospitalization, age ≥18 years old

The incidence rate according to time-varying exposure status is shown in Table 3. There was a total of 16 individuals with COVID-19-associated hospitalizations among those who received the 2024-2025 BNT162b2 COVID-19 vaccine and 1,120 among those who did not receive any 2024-2025 COVID-19 vaccine, corresponding to rates of COVID-19-associated hospitalization of 2.2 per 100,000 person-months (95% CI: 1.3-3.6) in the vaccinated group and 4.1 per 100,000 person-months (95% CI: 3.9-4.4) in the unvaccinated group. In adjusted models, overall BNT162b2 KP.2 VE (compared to not receiving a 2024-2025 COVID-19 vaccine of any kind regardless of prior vaccination history) was 41% (95% CI: 2-64%) against COVID-19-associated hospitalization **(Table 3)**.

**Table 3:** Rates of and Adjusted BNT162b2 COVID-19 Vaccine Effectiveness against COVID-19-Associated Hospital Admission Among Non-Immunocompromised Adults.

| Outcome and exposure status | Events<br>(person-months<br>at risk) | Incidence rate<br>per 100,000<br>person-months<br>(95% CI) | Adjusted VE<br>(95% CI) |
| --- | --- | --- | --- |
| *COVID-19-associated hospital admission, age ≥ 18 years* |  |  |  |
| Received 2024-2025 season BNT162b2 COVID-19 vaccine | 16<br>(719,540) | 2.2<br>(1.3 – 3.6) | 41%<br>(2 – 64) |
| Did not receive 2024-2025 COVID-19 vaccine of any kind | 1,120<br>(27,288,381) | 4.1<br>(3.9 – 4.4) | ref |
| *COVID-19-associated hospital admission, age ≥ 50 years* |  |  |  |
| Received 2024-2025 season BNT162b2 COVID-19 vaccine | 14<br>(459,439) | 3.0<br>(1.7 – 5.1) | 45%<br>(6 – 68) |
| Did not receive 2024-2025 COVID-19 vaccine of any kind | 933<br>(10,830,028) | 8.6<br>(8.1 – 9.2) | ref |
| *COVID-19-associated hospital admission, age ≥ 65 years* |  |  |  |
| Received 2024-2025 season BNT162b2 COVID-19 vaccine | 12<br>(230,842) | 5.2<br>(2.7 – 9.1) | 46%<br>(3 – 70) |
| Did not receive 2024-2025 COVID-19 vaccine of any kind | 705<br>(4,333,219) | 16.3<br>(15.1 – 17.5) | ref |

CI = confidence interval. Incidence rates are shown per 100,000 person-months at risk. Person-time at risk differ because outcomes were modelled independently, such that the occurrence of one did not interrupt time at risk for others.

### COVID-19-associated hospitalization, age ≥50 years old

Similar results were observed in the group of those aged ≥50 years old **(Table 3)**. Among those who had received a 2024-2025 COVID-19 vaccine in this group, there were 14 people with COVID-19-associated hospitalizations, corresponding to a rate of 3.0 hospitalizations per 100,000 person-months (95% CI: 1.7-5.1). Among those in this age group who had not received a 2024-2025 COVID-19 vaccine, there were 933 people with COVID-19-associated hospitalizations, corresponding to a rate of 8.6 per 100,000 person-months (95% CI: 8.1-9.2). In the adjusted model, BNT162b2 KP.2 VE for those aged ≥50 years old was 45% (95% CI: 6-68%) against COVID-19-associated hospitalization **(Table 3)**.

### COVID-19-associated outcomes, age ≥65 years old

This pattern held in the group of those aged ≥65 years old as well **(Table 3)**. Among those who had received a 2024-2025 COVID-19 vaccine in this group, there were 12 people with COVID-19-associated hospitalizations, corresponding to a rate of 5.2 hospitalizations per 100,000 person-months (95% CI: 2.7-9.1). Among those in this age group who had not received a 2024-2025 COVID-19 vaccine, there were 705 people with COVID-19-associated hospitalizations, corresponding to a rate of 16.3 per 100,000 person-months (95% CI: 15.1-17.5). Adjusted BNT162b2 KP.2 VE for those aged ≥65 years old was 46% (95% CI: 3-70%) against COVID-19-associated hospitalization **(Table 3)**.

## DISCUSSION

In this retrospective cohort study conducted between August 22, 2024, and December 31, 2024, among nearly 7 million non-immunocompromised adults in California and Louisiana, receipt of the BNT162b2 2024-2025 COVID-19 vaccine provided significant protection against serious COVID-19-associated hospitalization compared to those who did not receive a 2024-2025 season COVID-19 vaccine of any brand in this period. These results parallel the roughly 40% VE against COVID-19-associated hospitalization reported in other studies, most of which follow a test-negative rather than cohort design, underscoring the convergence of these findings.^4,15^

Of note, the majority of the people in both groups were at high risk of progression to severe disease (70% in the vaccinated group and 61% in the unvaccinated group) based on CDC high risk criteria.^11^ The outcome incidence rate as well as VE estimates in this study continue to demonstrate the value of an updated COVID-19 vaccines formulation that is well-matched to target the dominant circulating SARS-CoV-2 variant even in the context of high seropositive rates in the US population. Despite prior vaccination and/or natural infection, individuals are susceptible to subsequent infection due to waning immunity. Our findings underscore the need for universal revaccination as recommended by ACIP, as well as additional doses every six month for individuals ≥65 years of age.^16^ In addition, the findings support the June 5, 2024 FDA Vaccines and Related Biological Products Advisory Committee (VRBPAC) decision to update the 2024-2025 season COVID-19 vaccine formulation.^17^

Despite recommendations that all individuals age ≥6 months of age in the US receive a 2024-2025 COVID-19 vaccine, mid-season uptake of the BNT162b2 product among non-immunocompromised adults in California and Louisiana was only 4.7%. If adult 2024-2025 mid-season COVID-19 vaccine uptake were instead similar to mid-2024-2025 season influenza vaccine uptake (41.6% in adults as of December 28, 2024),^18^ and calculating the number needed to vaccinate as the reciprocal of the absolute risk difference, findings from this study suggest over 12,000 COVID-19-associated adult hospitalizations could have been avoided from August 22 – December 31, 2024 nationally. Those hospitalized incur significant medical costs, may experience critical illness, and, if discharged, are at higher risk of post-COVID conditions (“Long COVID”).^19^

There are various limitations associated with this analysis. In terms of generalizability, the individuals from California and Louisiana can be seen to represent a wide swath of the US population culturally and demographically. However, the two states alone may not truly represent the entire country in other important factors. This study utilizes claims data which limits us to relying on ICD-10 codes rather than laboratory-confirmed infection. Finally, these results describe the first four months of the 2024-2025 respiratory season and may not be representative of the entire season.^20^

## CONCLUSION

BNT162b2 2024-2025 COVID-19 vaccine provided significant protection against COVID-19-associated hospitalization for non-immunocompromised adults and was consistent among varying age strata. The findings support the utility of updated COVID-19 vaccine formulations that target contemporary circulating variants and the maintenance of routine vaccination recommendations.

## Data Availability

Pfizers data use agreement with HealthVerity prohibits sharing individual level data.

## ACKNOWLEDGEMENTS

All authors attest they meet the ICMJE criteria for authorship.

## Data Sharing Statement

Pfizer’s data use agreement with HealthVerity prohibits sharing individual-level data.

## Additional Contributions

The authors wish to thank Burcu Karakuzu Ikizler and Alison Randall for their contributions to study operations, and Xuezhe Wang for his contributions to data management and biostatistical modelling. All acknowledgees have reviewed the manuscript and provided written consent to be acknowledged.

## Data Access and Cleaning Methods

Several authors (KMA, JSM, TY, AZ) had access to the entire database population used to create the study population. Data cleaning methods and results were independently programmed by two programmers, with all discrepancies resolved. All authors had access to all summary data presented in this manuscript.

## REFERENCES

1. Panagiotakopoulos L. Use of COVID-19 Vaccines for Persons Aged ≥6 Months: Recommendations of the Advisory Committee on Immunization Practices — United States, 2024–2025. MMWR Morb Mortal Wkly Rep. 2024;73. doi:10.15585/mmwr.mm7337e2

2. Commissioner O of the. FDA Takes Action on Updated mRNA COVID-19 Vaccines to Better Protect Against Currently Circulating Variants. FDA. September 11, 2023. Accessed March 20, 2024. https://www.fda.gov/news-events/press-announcements/fda-takes-action-updated-mrna-covid-19-vaccines-better-protect-against-currently-circulating

3. Office of the Commissioner. FDA Approves and Authorizes Updated mRNA COVID-19 Vaccines to Better Protect Against Currently Circulating Variants. FDA. August 23, 2024. Accessed May 8, 2025. https://www.fda.gov/news-events/press-announcements/fda-approves-and-authorizes-updated-mrna-covid-19-vaccines-better-protect-against-currently

4. Appaneal HJ, Lopes VV, Puzniak L, et al. Early effectiveness of the BNT162b2 KP.2 vaccine against COVID-19 in the US Veterans Affairs Healthcare System. Nat Commun. 2025;16(1):4033. doi:10.1038/s41467-025-59344-7

5. Link-Gelles R. Interim Estimates of 2024–2025 COVID-19 Vaccine Effectiveness Among Adults Aged ≥18 Years — VISION and IVY Networks, September 2024– January 2025. MMWR Morb Mortal Wkly Rep. 2025;74. doi:10.15585/mmwr.mm7406a1

6. Andersen KM, Allen KE, Nepal RM, et al. Effectiveness of BNT162b2 XBB.1.5 vaccine in immunocompetent adults using tokenization in two U.S. states. Vaccine. 2025;52:126881. doi:10.1016/j.vaccine.2025.126881

7. McGrath LJ, Khan FL, Cook AD, et al. Tokenization and Linkage of State Vaccine Registry Data to Improve Determination of COVID-19 Vaccination Status in a Large Administrative Claims Database. Pharmacoepidemiol Drug Saf. 2024;33(S2):e5891. doi:10.1002/pds.5891

8. McGrath LJ, Khan FL, Lopez SMC, et al. 2023-2024 COVID-19 vaccine uptake among immunocompromised individuals in two US states. Vaccine. 2025;56:127120. doi:10.1016/j.vaccine.2025.127120

9. Rubin LG, Levin MJ, Ljungman P, et al. 2013 IDSA clinical practice guideline for vaccination of the immunocompromised host. Clin Infect Dis Off Publ Infect Dis Soc Am. 2014;58(3):309–318. doi:10.1093/cid/cit816

10. Andersen KM, Brouillette MA, Mateus JS, et al. Distinguishing Hospital Encounters as “with” vs “for” COVID-19 Using A Large US Administrative Claims Database. Pharmacoepidemiol Drug Saf. 2024;33(S2):e5891.

11. CDC. People with Certain Medical Conditions. Centers for Disease Control and Prevention. May 11, 2023. Accessed March 20, 2024. https://www.cdc.gov/coronavirus/2019-ncov/need-extra-precautions/people-with-medical-conditions.html

12. Wiemken TL, McGrath LJ, Andersen KM, et al. Coronavirus Disease 2019 Severity and Risk of Subsequent Cardiovascular Events. Clin Infect Dis. Published online August 19, 2022:ciac661. doi:10.1093/cid/ciac661

13. McGrath LJ, Malhotra D, Miles AC, et al. Estimated Effectiveness of Coadministration of the BNT162b2 BA.4/5 COVID-19 Vaccine With Influenza Vaccine. JAMA Netw Open. 2023;6(11):e2342151. doi:10.1001/jamanetworkopen.2023.42151

14. Efron B. The Efficiency of Cox’s Likelihood Function for Censored Data. J Am Stat Assoc. 1977;72(359):557–565. doi:10.2307/2286217

15. Rudolph AE, Khan FL, Sun X, et al. Receipt of BNT162b2 KP.2 Vaccine and COVID-19 CVS MinuteClinic Visits in US Adults. Published online January 17, 2025:2025.01.15.24319342. doi:10.1101/2025.01.15.24319342

16. Centers for Disease Control and Prevention. SARS-CoV-2 Variant Proportions. Laboratory Surveillance. July 5, 2024. https://data.cdc.gov/Laboratory-Surveillance/SARS-CoV-2-Variant-Proportions/jr58-6ysp/about_data

17. Center for Biologics Evaluation and Research (CBER). Summary Minutes from 185th Vaccines and Related Biological Products Advisory Committee (VRBPAC). June 5, 2024. https://www.fda.gov/media/180732/download

18. CDC. Influenza Vaccination Coverage and Intent for Vaccination, Adults 18 Years and Older, United States. FluVaxView. May 15, 2025. Accessed May 19, 2025. https://www.cdc.gov/fluvaxview/dashboard/adult-coverage.html

19. Pierre V, Draica, Florin, Di Fusco, Manuela, et al. The impact of vaccination and outpatient treatment on the economic burden of Covid-19 in the United States omicron era: a systematic literature review. J Med Econ. 2023;26(1):1519–1531. doi:10.1080/13696998.2023.2281882

20. Bartsch SM, Chin KL, Strych U, et al. The Current and Future Burden of Long COVID in the United States (U.S.). J Infect Dis. Published online January 22, 2025:jiaf030. doi:10.1093/infdis/jiaf030

